# Novel risk factors for Coronavirus disease-associated mucormycosis (CAM): a case control study during the outbreak in India

**DOI:** 10.1101/2021.07.24.21261040

**Authors:** Umang Arora, Megha Priyadarshi, Varidh Katiyar, Manish Soneja, Prerna Garg, Ishan Gupta, Vishwesh Bharadiya, Parul Berry, Tamoghna Ghosh, Lajjaben Patel, Radhika Sarda, Shreya Garg, Shubham Agarwal, Veronica Arora, Aishwarya Ramprasad, Amit Kumar, Rohit Kumar Garg, Parul Kodan, Neeraj Nischal, Gagandeep Singh, Pankaj Jorwal, Arvind Kumar, Upendra Baitha, Ved Prakash Meena, Animesh Ray, Prayas Sethi, Immaculata Xess, Naval Vikram, Sanjeev Sinha, Ashutosh Biswas, Alok Thakar, Sushma Bhatnagar, Anjan Trikha, Naveet Wig

## Abstract

**Background:** The epidemiology of the Coronavirus-disease associated mucormycosis (CAM) syndemic is poorly elucidated. We aimed to identify risk factors that may explain the burden of cases and help develop preventive strategies.

**Methods:** We performed a case-control study comparing cases diagnosed with CAM and those who had recovered from COVID-19 without developing mucormycosis (controls). Information on comorbidities, glycemic control, and practices related to COVID-19 prevention and treatment was recorded.

**Results:** 352 patients (152 cases and 200 controls) diagnosed with COVID-19 during April-May 2021 were included. In the CAM group, symptoms of mucormycosis began a mean 18.9 (SD 9.1) days after onset of COVID-19, and predominantly rhino-sinus and orbital involvement was present. All, but one, CAM cases carried conventional risk factors of diabetes and steroid use. On multivariable regression, increased odds of CAM were associated with the presence of diabetes (adjusted OR 3.5, 95%CI 1.1-11), use of systemic steroids (aOR 7.7,95% CI 2.4-24.7), prolonged use of cloth and surgical masks (vs no mask, aOR 6.9, 95%CI 1.5-33.1), and repeated nasopharyngeal swab testing during the COVID-19 illness (aOR 1.6,95% CI 1.2-2.2). Zinc therapy, probably due to its utility in immune function, was found to be protective (aOR 0.05, 95%CI 0.01-0.19). Notably, the requirement of oxygen supplementation or hospitalization did not affect the risk of CAM.

**Conclusion:** Judicious use of steroids and stringent glycemic control are vital to preventing mucormycosis. Use of clean masks, preference for N95 masks if available, and minimizing swab testing after the diagnosis of COVID-19 may further reduce the incidence of CAM.

## Introduction

A recent upsurge in the cases of mucormycosis has been evident during the waxing and waning course of the COVID-19 pandemic, resulting in a syndemic named the Coronavirus diseases-associated mucormycosis (CAM).^1^

Mucormycosis is an opportunistic disease caused by angio-invasive fungi of the order *Mucorales*, with an estimated prevalence of 140 cases per million population in India.^2^ It is characterized by the rapid development of tissue necrosis manifesting as rhino-orbital-cerebral, pulmonary, cutaneous, gastrointestinal or disseminated disease. It is almost exclusively seen in patients having immunocompromised status due to diabetes, malignancy, chemotherapy, or immunosuppressive drugs.^2^

A multicentric cohort from India reported a two-fold rise in the prevalence of mucormycosis between September–December 2020 compared to the same time period in 2019 as a consequence of the increased burden of CAM.^1^ Moreover, the number of cases rose even higher during the period of April-May 2021.^3^ High rates of morbidity and mortality, and exorbitant out-of-pocket expenditure for antifungal therapy with liposomal amphotericin B, posaconazole, and isavuconazole have made it imperative to prevent the occurrence of this disease.^4^

CAM occurs during or soon after convalescence from COVID-19. The epidemiological link with the COVID-19 infection suggests that novel non-conventional risk factors may have a role in CAM infection. Hypotheses abound in medical literature based on information available from mucormycosis prior to the onset of COVID-19, particularly existing knowledge of factors required for growth or inhibition of *Mucorales* in-vitro. We performed a case-control study comparing CAM patients with patients who have recovered from COVID-19 without developing mucormycosis, to investigate putative risk factors that may have contributed to the development of CAM.

## Methods

A case-control study was performed over a period of 15 days from 1^st^ to 15^th^ June 2021 at a tertiary care center in Delhi, India. Patients were selected either as cases of Coronavirus disease-associated mucormycosis or controls who had recovered from COVID-19 without developing mucormycosis. Informed consent was obtained from patients and the study was approved by the Institute Ethics Committee.[IECPG-353/28.05.21]

Patients above 18 years of age diagnosed with CAM were identified as cases. CAM was defined as mucormycosis based on clinico-radiological or microbiological evidence with fungal staining or culture. As per the consensus criteria, definite mucormycosis was defined as demonstration of aseptate ribbon-like broad hyphae on histopathology of biopsied samples (with or without culture) and probable mucormycosis as patients with suggestive clinical symptoms and radiologic imaging along with isolation of *Mucorales* from nonsterile samples such as swab sample from nasal area.^4^ Controls were defined as patients who recovered from COVID-19 with symptom onset at least one month prior to recruitment and no clinical features suggestive of CAM. Controls were selected consecutively from records of ward as well as out-patient clinic. The case definition by World Health Organization(WHO) for definite or probable cases of SARS-CoV-2 infection was used to select both cases and controls.^5^ We limited both cases and controls to those who developed symptoms or test positivity of COVID-19 illness during the months of April–May 2021 to account for potential confounding factors including change in the viral strains. Patients who did not give consent were excluded.

We assessed the risk factors associated with the development of mucormycosis in patients with COVID-19 using a structured form which included relevant information regarding demographic details, comorbidities, glycemic control (poor glycemic control defined as blood glucose ≥200 mg/dL), COVID-19 vaccination status, COVID-19 related practices (mask-wearing, steam inhalation), COVID-19 infection severity as per WHO guidelines (oxygen or hospitalization requirement), and treatment given during COVID-19 illness particularly steroid therapy. Cases were interviewed during their admission while controls were contacted telephonically. An investigator-guided online form and digital consent was used for discharged patients. Missing information was supplemented using telephonic contact with patients’ family members, discharge records, prescription slips, and hospital charts if needed. HbA1c and ferritin levels, if checked during management of COVID-19, were recorded. Doses of various steroids (methylprednisolone, prednisone) were converted to equivalent doses of dexamethasone.^6^

Categorical variables were expressed as frequency and proportion of patients in the two groups were compared using the chi-square test. Continuous variables were checked for approximate normality using the Shapiro Wilk test. Normally distributed variables were expressed as mean (SD) and compared using unpaired t-test, otherwise median (IQR) were presented and compared using Wilcoxon rank-sum test. A p-value of less than 0.05 was considered statistically significant. Univariate logistic regression for all potential risk factors was performed for the occurrence of mucormycosis in patients recovered from COVID-19. Variables with a p-value of less than 0.2 as well as known risk factors for mucormycosis were included in the multivariable model. Multivariable logistic regression analysis was performed with backward elimination to develop the final regression model. Statistical analysis was performed using Stata version 14 (TX, USA) and graphical representation using R version 4.0.2 and *RStudio* 1.4.1717.

## Results

A total of 163 cases of CAM were screened for inclusion. Eleven patients were excluded; seven did not give consent, three were diagnosed with *Aspergillus* on fungal culture, and one patient had suffered from COVID-19 more than six months before the diagnosis of mucormycosis. Finally, 152 patients of CAM (cases) and 200 patients of COVID-19 without mucormycosis (controls) were included in the study. The spectrum of involvement of CAM cases included rhino-sinus (n=44, 29%), rhino-orbital (n=72, 47.3%), rhino-orbito-cerebral (n=22, 14.5%), isolated orbital (n=2, 1.3%), rhino-orbital and palatal (n=8, 5.3%), cutaneous (n=1, 0.6%), isolated pulmonary (n=2, 1.3%), and disseminated with rhino-sinus and pulmonary involvement (n=1, 0.6%). Cases were interviewed at a median of 46 days (Range: 30 – 70), and controls 48 days (Range: 30 –72) after onset of COVID-19 symptoms. The time period when COVID-19 symptoms began were distributed over April–May 2021 in both groups.(Figure 1) In patients with CAM, symptoms of mucormycosis began at a mean of 18.9 ± 9.1 (Range: 2–47) days after symptom onset of COVID-19. (Figure 2) The mean age of the study population was 48.2 ± 14 years with 226 males (64.2%). Age, sex, and body mass index (BMI) were similar in the two groups. A greater proportion of CAM cases resided in rural areas as compared to controls (p <0.001). (Table 1)

**Table 1:**
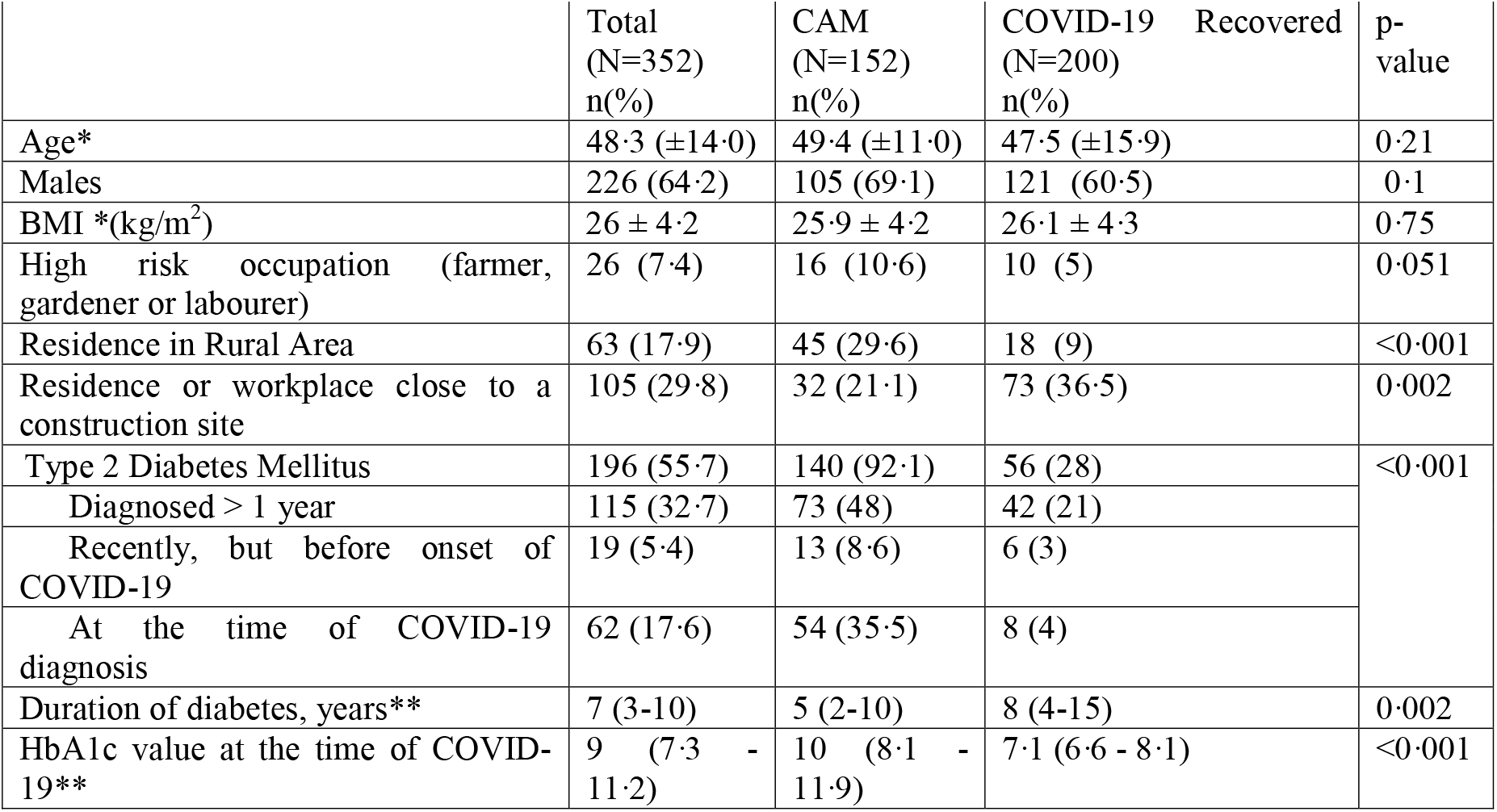
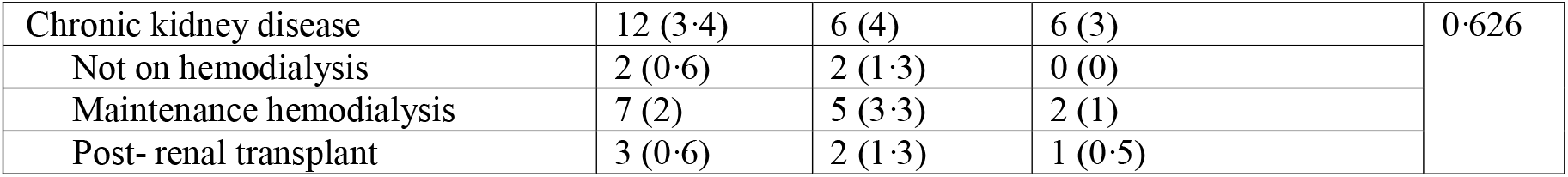
Demographic and comorbidity profiles of cases compared to controls. All values represented as n(%) except for *mean ±SD and **median (IQR)

|  | Total<br>(N=352)<br>n(%) | CAM<br>(N=152)<br>n(%) | COVID-19 Recovered<br>(N=200)<br>n(%) | p-value |
| --- | --- | --- | --- | --- |
| Age* | 48.3 (±14.0) | 49.4 (±11.0) | 47.5 (±15.9) | 0.21 |
| Males | 226 (64.2) | 105 (69.1) | 121 (60.5) | 0.1 |
| BMI *(kg/m<sup>2</sup>) | 26 ± 4.2 | 25.9 ± 4.2 | 26.1 ± 4.3 | 0.75 |
| High risk occupation (farmer, gardener or labourer) | 26 (7.4) | 16 (10.6) | 10 (5) | 0.051 |
| Residence in Rural Area | 63 (17.9) | 45 (29.6) | 18 (9) | <0.001 |
| Residence or workplace close to a construction site | 105 (29.8) | 32 (21.1) | 73 (36.5) | 0.002 |
| Type 2 Diabetes Mellitus | 196 (55.7) | 140 (92.1) | 56 (28) | <0.001 |
| Diagnosed > 1 year | 115 (32.7) | 73 (48) | 42 (21) |  |
| Recently, but before onset of COVID-19 | 19 (5.4) | 13 (8.6) | 6 (3) |  |
| At the time of COVID-19 diagnosis | 62 (17.6) | 54 (35.5) | 8 (4) |  |
| Duration of diabetes, years** | 7 (3-10) | 5 (2-10) | 8 (4-15) | 0.002 |
| HbA1c value at the time of COVID-19** | 9 (7.3 - 11.2) | 10 (8.1 - 11.9) | 7.1 (6.6 - 8.1) | <0.001 |
| Chronic kidney disease | 12 (3.4) | 6 (4) | 6 (3) | 0.626 |
| Not on hemodialysis | 2 (0.6) | 2 (1.3) | 0 (0) |  |
| Maintenance hemodialysis | 7 (2) | 5 (3.3) | 2 (1) |  |
| Post- renal transplant | 3 (0.6) | 2 (1.3) | 1 (0.5) |  |

**Figure 1:**
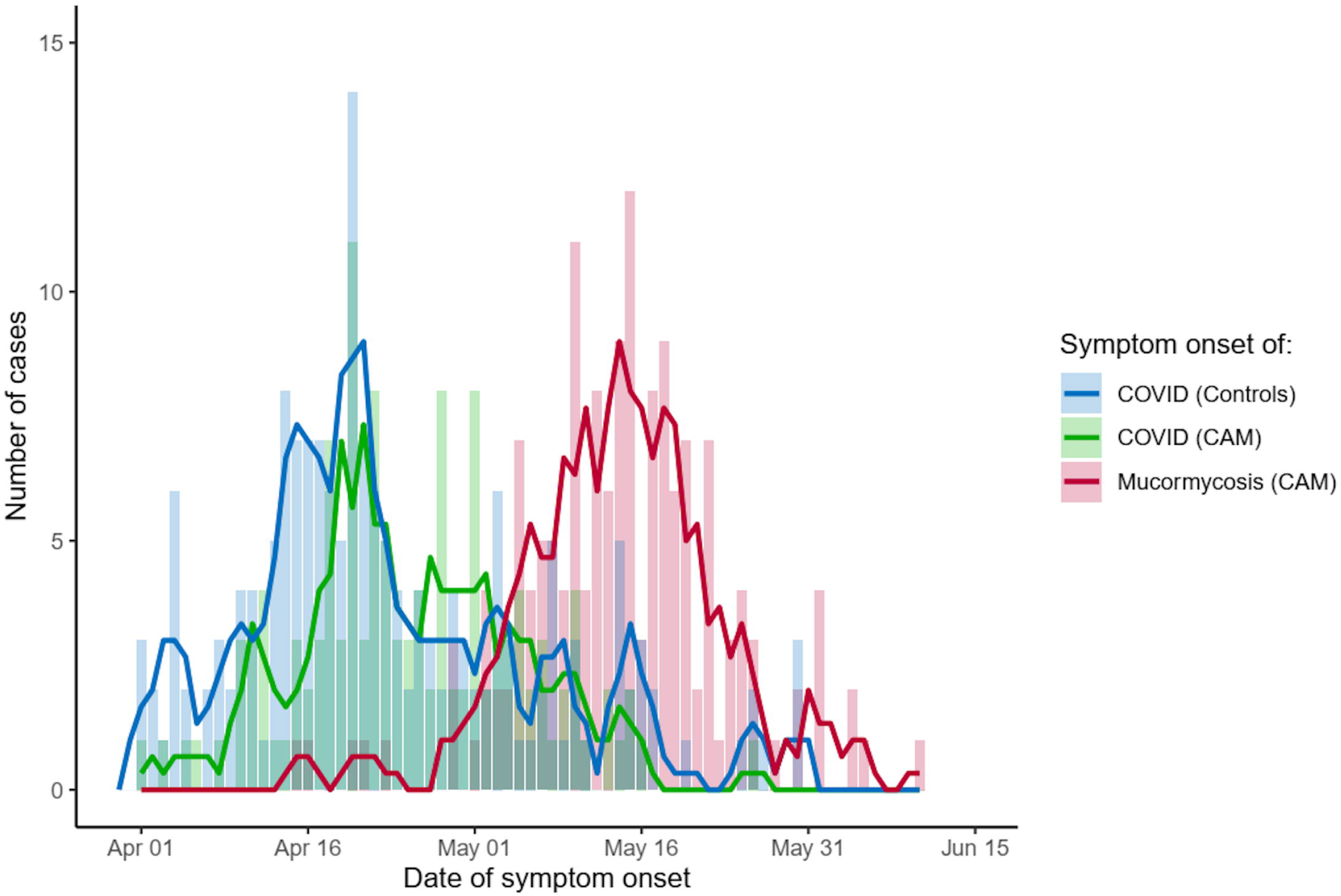
Date of symptom onset of COVID-19 illness for cases (green) and controls (blue), along with date of onset of mucormycosis symptoms for consecutive cases of CAM (red). The peak of symptom onset of COVID-19 for both groups was 20 April 2021 and of mucormycosis for cases was 15 May 2021.

**Figure 2:**
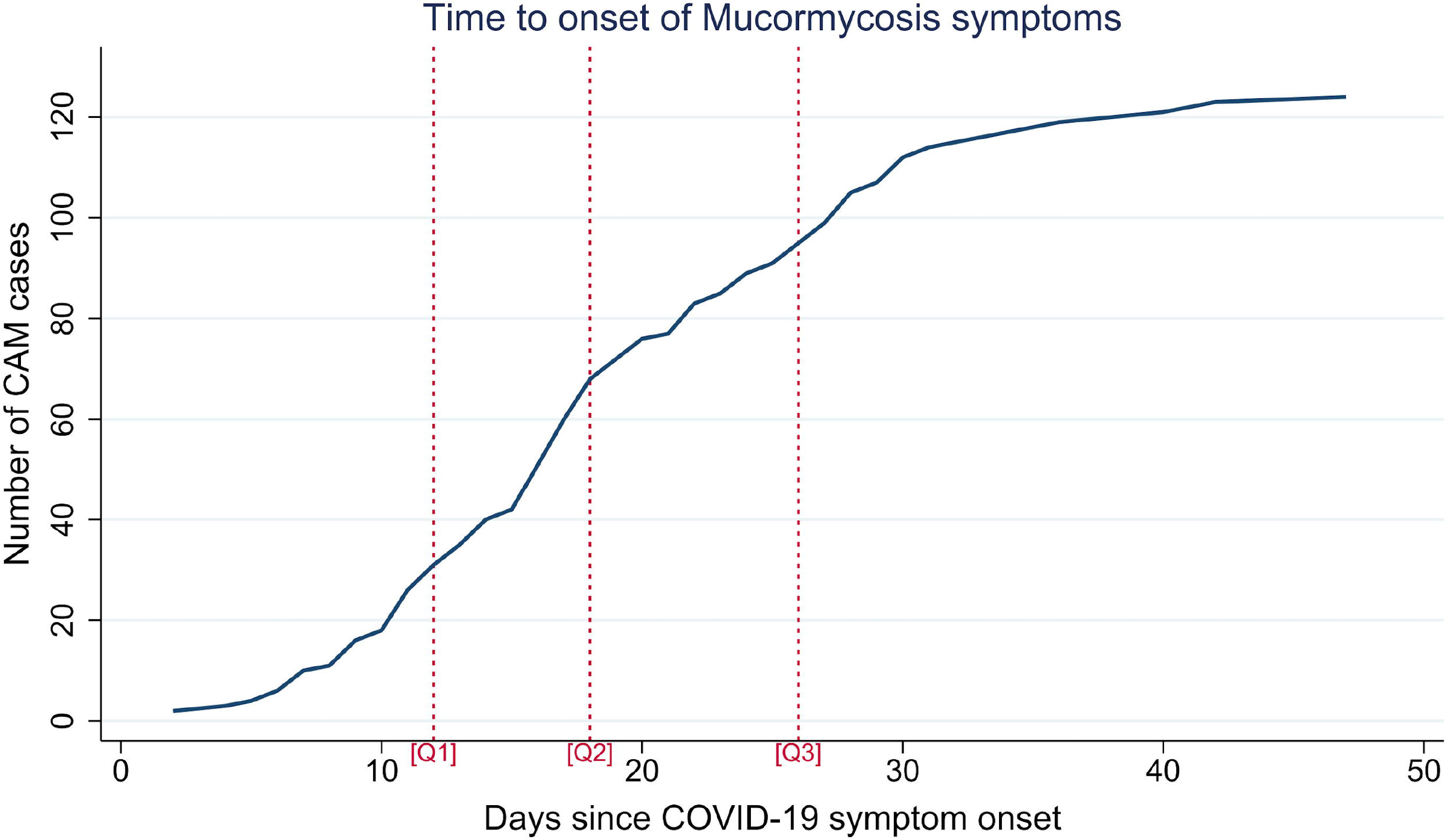
Cumulative time to symptom onset of mucormycosis after the onset of COVID-19 symptoms in CAM. The first (Q1), second (Q2) and third quartile (Q3) represent 12, 18 and 26 days respectively. CAM patients who were asymptomatic for COVID-19 are not included (n=29).

The most frequent comorbidities seen in our study population were diabetes (n=196, 55.7%), hypertension (n=125, 35.3%) and chronic kidney disease (n=12, 3.4%). Diabetes was significantly more frequent among cases than controls (92.1% vs 28%, p <0.001). A third of the diabetic patients with CAM (n=54, 35.5%) were newly diagnosed with diabetes during hospitalization for COVID-19 based on elevated blood glucose or abnormal HbA1c levels. None of our patients had malignancy or were on chemotherapy.

Most of the patients in both groups had mild COVID-19. (Table 2) The requirement of hospitalization and oxygen therapy was similar in both groups. Patients with severe COVID-19 were more common amongst the controls (21% vs 9.9%, p<0.001). Ferritin levels during acute phase of COVID-19 disease were available for 174 patients, which were higher in the CAM group (p<0.001). More than half of the study population received systemic steroids for the management of COVID-19, being more frequently used in CAM cases than controls (65.8% vs 48%,p=0.001). Zinc use was more common in the control group (79.9% vs 53.8%, p<0.001). Poor glycemic control was observed in a higher proportion of cases than controls (90.6% vs 51.5%, p <0.001). Information on the occurrence of diabetic ketoacidosis was available for 176 patients. Six patients presented with DKA, and all belonged to the CAM group (5%). The overall distribution of patients between the two groups, according to the best established risk factors diabetes and systemic steroids is depicted in Figure 3.

**Table 2:**
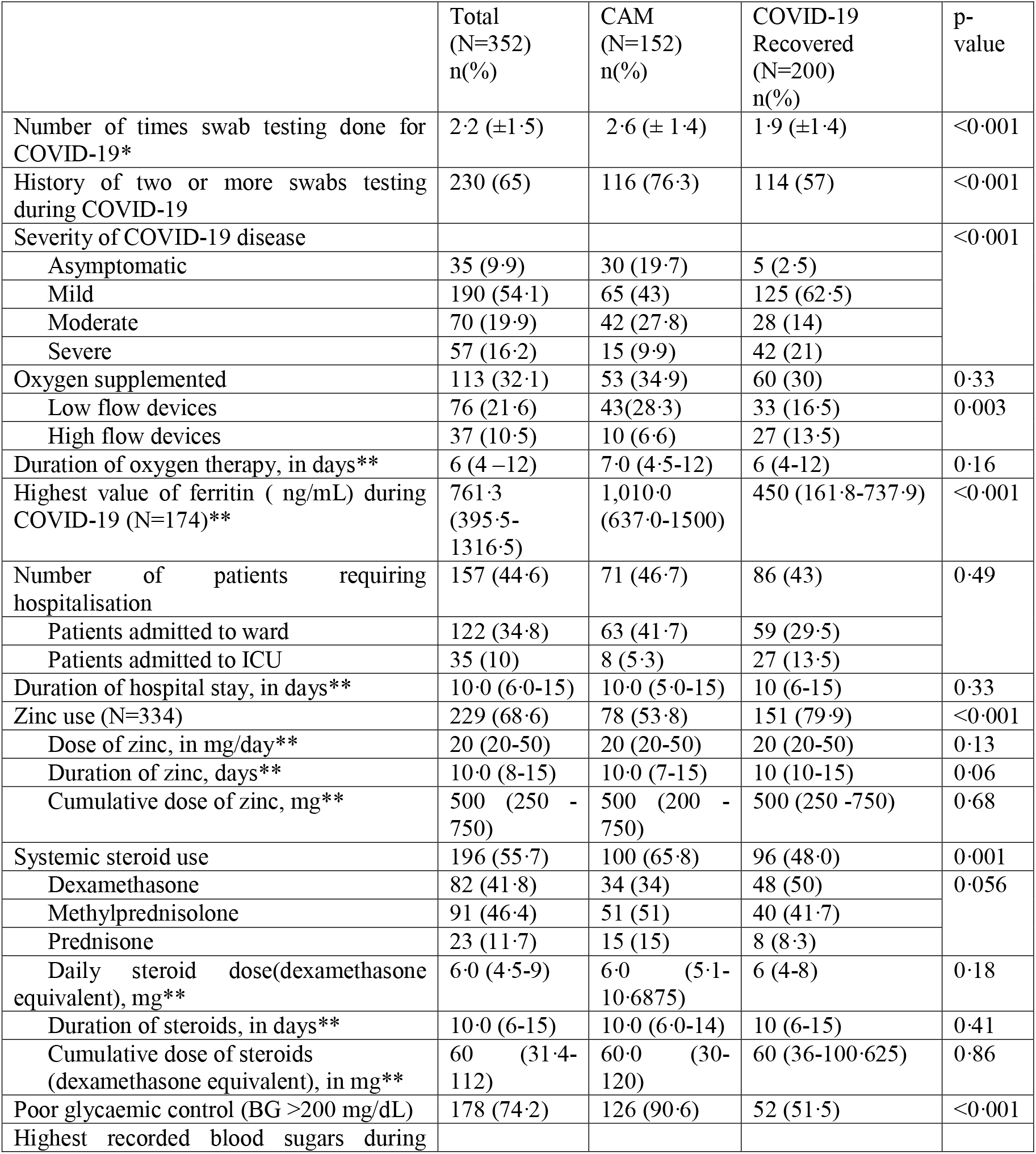
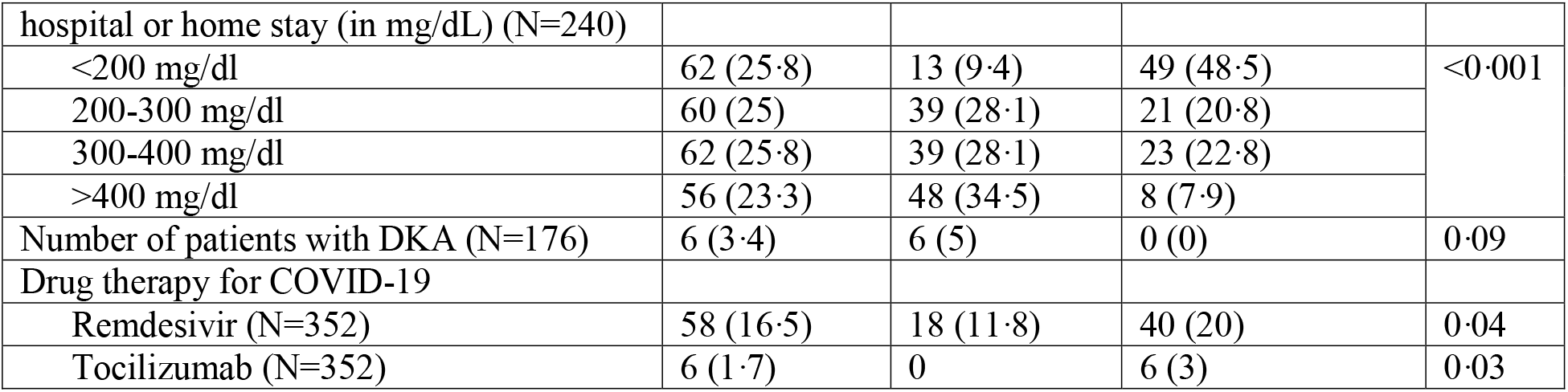
Diagnosis, disease severity, and management of COVID-19 illness in cases compared to controls. All values represented as n(%) except for *mean ±SD and **median (IQR)

**Figure 3:**
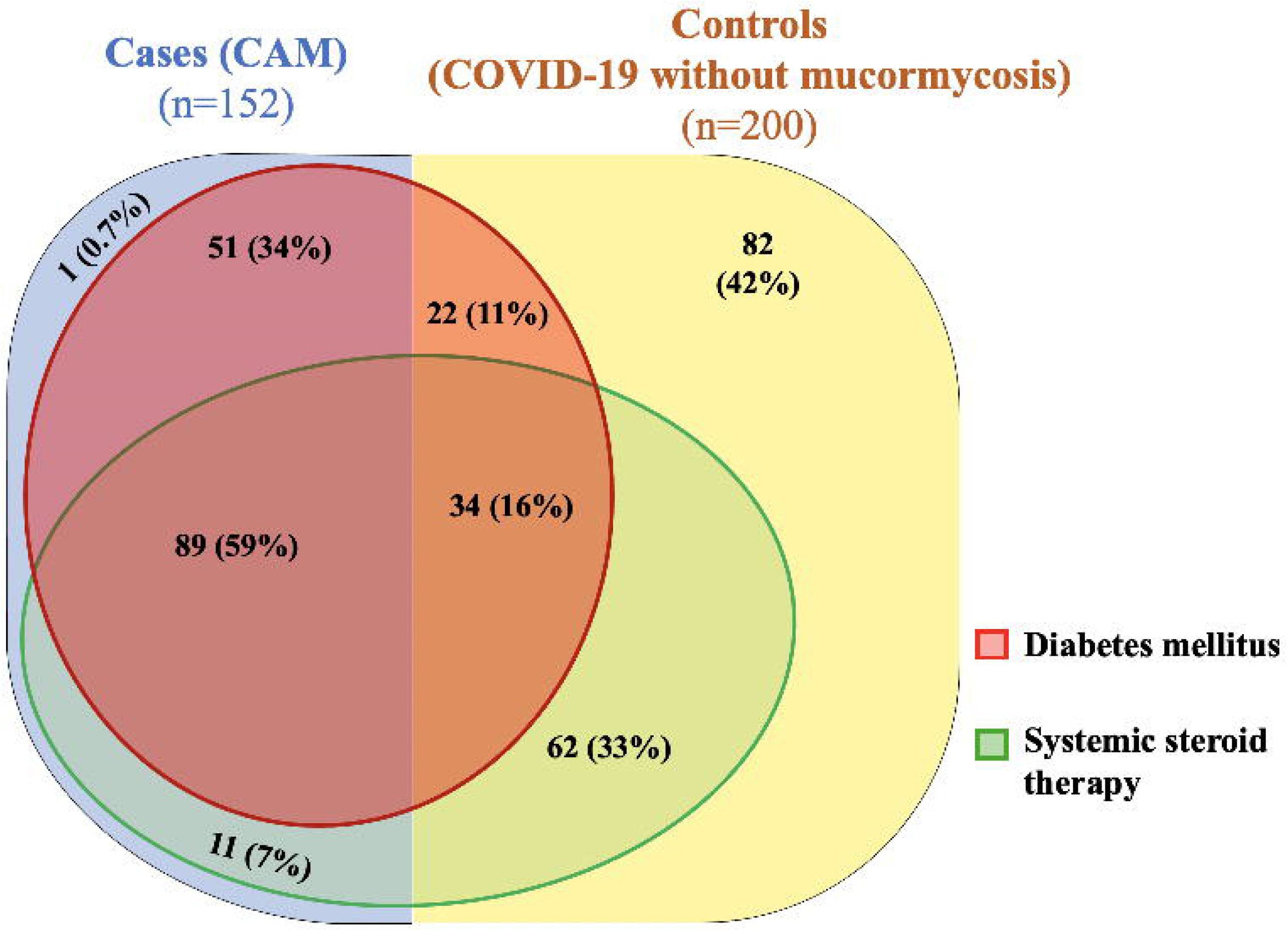
Venn diagram depicting the distribution of cases with coronavirus disease-associated mucormycosis (CAM, blue) and controls (COVID-19 cases recovered without mucormycosis, yellow) in relation to the two main risk factors (diabetes mellitus, red; and systemic steroid therapy, green). Percentage out of the cases and controls are represented for each region in the parentheses.

N95 mask use was more frequent among controls (42.5% vs 18%), while CAM patients tended to use cloth masks (52% vs 36%). (Table 3) Use of N95 masks was observed to be protective in comparison to not using a mask (p<0.05). Use of cloth masks more than four hours (4–6 hours, p=0.002; >6 hours, p<0.0001) and use of surgical masks for more than six hours (p=0.002) was found to be associated with a higher risk of CAM when compared to the use of respective masks for less than two hours or N95 for any duration. (Figure 4) The findings were similar even after excluding health care workers (n=40) as confounders, who were more limited to the control group and used N95 masks exclusively. Due to diversity in practices of mask disinfection, reuse, and discarding, this information could not be analyzed. The two groups did not differ in their practice of steam inhalation vis-a-vis the frequency of steam inhalation and the time of starting steam inhalation in relation to the onset of COVID-19 symptoms.

**Table 3:** Mask use and steam inhalation practices in cases vs controls. All values represented as n(%)

|  | Total (N=352)<br>n(%) | CAM (N=152)<br>n(%) | COVID-19<br>Recovered<br>(N=200), n(%) | p-value |
| --- | --- | --- | --- | --- |
| Type of mask used before diagnosis of<br>COVID-19 (for most days in the<br>preceding month) (N=350) |  |  |  | < 0.001 |
| N95 mask | 112 (32) | 27 (18) | 85 (42.5) |  |
| Surgical mask | 58 (16.6) | 29 (19.3) | 29 (14.5) |  |
| Cloth mask | 150 (42.9) | 78 (52) | 72 (36) |  |
| None | 30 (8.6) | 16 (10.7) | 14 (7) |  |
| Duration of mask use per day(N=319) |  |  |  | 0.037 |
| <2 hours | 101 (31.7) | 33 (24.6) | 68 (36.7) |  |
| 2-4 hours | 83 (26) | 31 (23.1) | 52 (28.1) |  |
| 4-6 hours | 39 (12.2) | 19 (14.2) | 20 (10.8) |  |
| >6 hours | 96 (30.1) | 51 (37.4) | 45 (24.3) |  |
| Frequency of steam inhalation |  |  |  | 0.98 |
| Two or more times per day | 191 (71.5) | 78 (72.2) | 113 (71.1) |  |
| Once a day | 41 (15.4) | 16 (14.8) | 25 (15.7) |  |
| Infrequently | 35 (13.1) | 14 (13) | 21 (13.2) |  |
| Timing of initiation of steam inhalation<br>with respect to COVID-19 illness<br>(N=261) |  |  |  | 0.98 |
| Since before symptoms of COVID-19 | 71 (27.2) | 28 (27.7) | 43 (26.9) |  |
| During COVID-19 illness alone | 175 (67) | 67 (66.3) | 108 (67.5) |  |
| Continued after recovery from<br>COVID-19 | 15 (5.7) | 6 (5.9) | 9 (5.6) |  |

**Figure 4:**
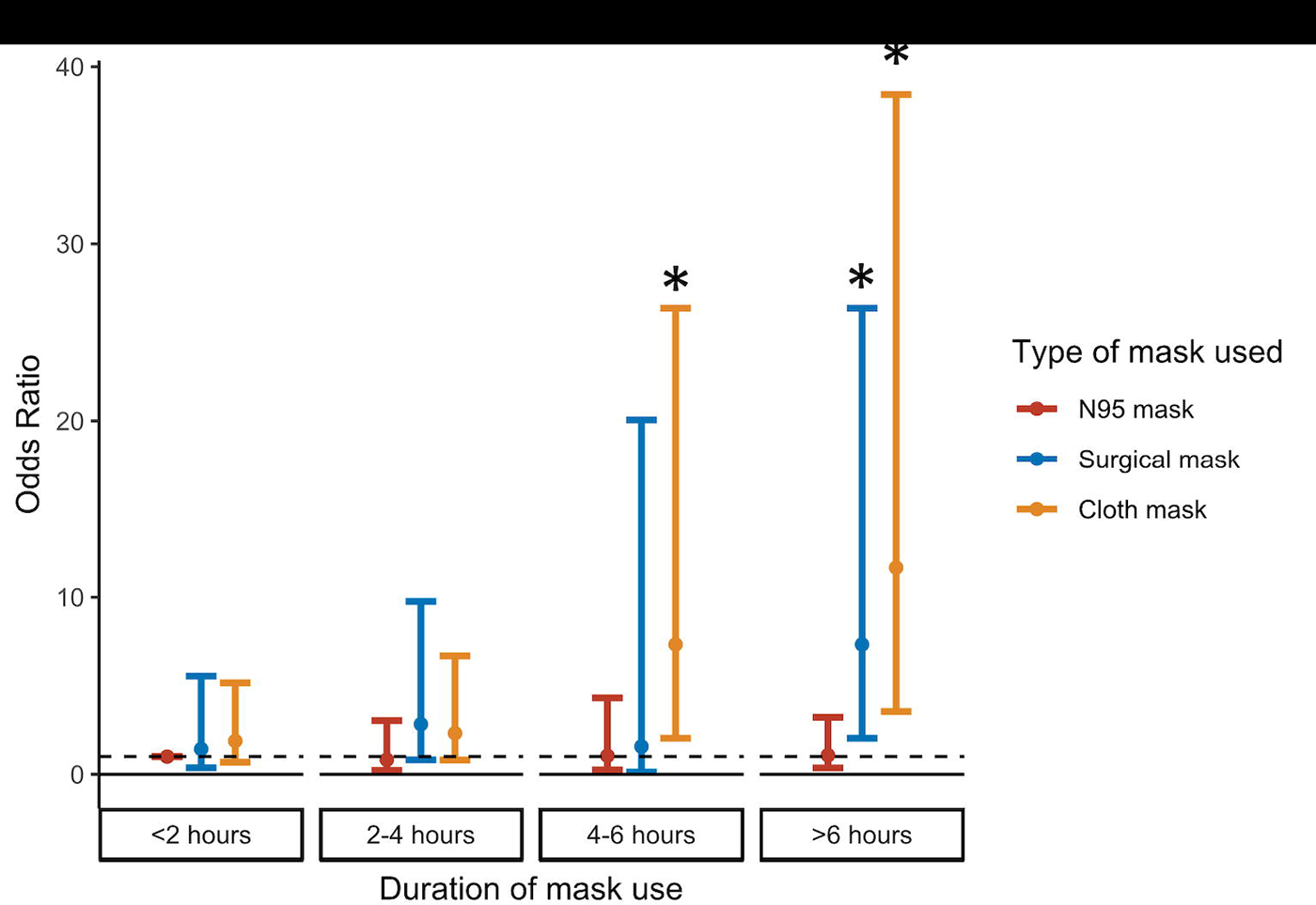
Risk of Coronavirus disease-associated mucormycosis depends on the type of mask as well as duration of usage. * represents significant difference compared to use of N95 mask for <2 hours

Mask use practices were divided into four risk categories based on the univariate analysis. Use of N95 masks for any duration was considered as low risk, while use of no mask was categorized separately. Use of surgical masks for more than 6 hours or cloth masks for more than 4 hours constituted the third group with highest risk, while shorter duration of either mask use(surgical mask <6 hours or cloth masks <4 hours) formed the fourth group. The multivariable model was limited to patients with symptomatic COVID-19 since hospitalization policy excluded admission of asymptomatic COVID-19 patients. Risk factors that were found to be independently associated with the risk of development of mucormycosis in multivariable analysis were diabetes, mild and moderate (vs severe) COVID-19 disease, highest blood glucose of more than 200 mg/dL at any time during COVID-19 illness, the number of swab tests performed, and systemic steroid use.(Table 4) The use of zinc was found to be protective. Among mask use, prolonged use of surgical masks (>6 hours) and cloth masks (>4 hours) were associated with increased odds of CAM (6.9, 95% CI 1.5 – 33.1, p=0.02) compared to no mask use. Shorter duration of cloth or surgical mask usage was associated with higher odds of CAM vs N95 masks (4.9, 95% CI 1.9 – 16, p<0.001), but lower than prolonged surgical or cloth mask use (0.28, 95% CI 0.08 – 0.97, p=0.046). The findings were consistent after subgroup analysis of the same mask use categories among patients residing in urban areas.

**Table 4:** Multivariable analysis of risk factors for development of Coronavirus disease-associated mucormycosis.

| **Risk factor** | **Adjusted odds ratio (95% CI)** | **P-value** |
| --- | --- | --- |
| Type 2 Diabetes mellitus | 3.5 (1.1–11) | 0.032 |
| Steroid use | 7.7 (2.4–24.7) | 0.001 |
| Severity of COVID-19 disease (vs mild) |  |  |
| Moderate | 1.4 (0.4–4.2) | 0.58 |
| Severe | 0.17 (0.05–0.56) | 0.004 |
| Highest recorded blood glucose levels (vs $\leq 200$ mg/dL) | | |
| 200-299 mg/dL | 5.4 (1.2–24.8) | 0.03 |
| 300-399 mg/dL | 10.3 (2.7–46.6) | 0.03 |
| $\geq 400$ mg/dL | 27.0 (5.3–136.1) | <0.001 |
| Number of swab tests prior to onset of mucormycosis | 1.6 (1.2–2.2) | 0.001 |
| Zinc use | 0.05 (0.01–0.19) | <0.001 |
| Mask use (vs no mask use) |  |  |
| N95 mask use (any duration) | 0.39 (0.94 - 1.66) | 0.21 |
| Short duration use of surgical (<6 hours) or cloth mask (<4 hours) | 1.96 (0.48-8.06) | 0.35 |
| Prolonged use of surgical (>6 hours) or cloth mask (>4 hours) | 6.9 (1.5-33.1) | 0.02 |

## Discussion

Mucormycosis is a sporadic disease occurring almost exclusively in immunocompromised patients.^7^ The sudden spike in the incidence of CAM in the wake of the COVID-19 pandemic raises the possibility that COVID-19 infection may itself predispose to mucormycosis. This may occur directly through its impact on immune system or indirectly due to the interventions related to COVID-19 prevention and management.

Even in the pre-COVID-19 era, India reported the highest prevalence of mucormycosis worldwide, nearly 70 times that of global estimates.^2^ This disproportionate burden has been attributed to the high prevalence of both patient factors (diabetes) and environmental factors (hot humid environment).^8^ Residence in a rural area was more common among CAM cases, possibly because of a referral bias for a difficult to treat disease. Our study found that CAM was more common in males in the fifth and sixth decades. The disease occurred three weeks after onset of COVID-19 symptoms and involved mainly the rhino-orbital areas, similar to reported literature.^1^ Poor glycemic control, systemic steroid therapy, and presence of diabetes are associated with increased risk of CAM.^8^ However, 2-32% of reported cases of CAM may lack any of these associations.^1,9^ This is a departure from our finding, where a single case of CAM (out of 152) was neither diabetic nor had received steroids.

### Immune system modulation due to COVID-19, diabetes, and steroids

Neutrophils and macrophages kill sporangiospores and hyphal forms of *Mucorales* and constitute the major barrier to invasion.^10^ An adaptive immune response is stimulated characterized by strong Th-17 activation that initiates a stronger neutrophil response. Simultaneously, the immune response to COVID-19 is complex. Neutrophils in the nasopharyngeal epithelium demonstrate markers of premature activation,^11^ while the cells of adaptive immunity (T cells, NK cells, and B cells) are reduced in numbers.^12^ These abnormalities tend to peak in the second week, which may explain the clustering of cases of CAM in the third week after COVID-19 symptom onset. Also, endothelial dysfunction and vasculopathy due to COVID-19 may support angioinvasion and spread of *Mucorales*.^13^ Influence of the mutated variants of COVID-19 on the development of CAM remains to be explored.

Diabetes or impaired glucose tolerance is reported in the majority of patients with mucormycosis with or without COVID-19. Hyperglycemia inhibits neutrophil chemotaxis, phagocytosis by macrophages and degranulation of NK cells.^14^ It promotes surface glucose-regulated protein (GRP78) expression on the endothelium which is essential for invasion by *Mucorales*.^15^ The prevalence of diabetes in CAM was higher than historical cohorts of mucormycosis not associated with COVID (92% vs 76%).^16^ Newly detected diabetes is reportedly more prevalent in CAM compared to mucormycosis not associated with COVID-19 (21% vs 10%, p=0.02).^1^ However, these studies did not report measuring HbA1c for their mucormycosis patients, which may have led to the underreporting of diabetes in previous cohorts. Additionally, supra-physiological stress during COVID-19 illness and viral-mediated islet cell damage may contribute to hyperglycemia.^17^ Diabetic ketoacidosis is detected in 8–22% of mucormycosis patients at presentation, being a rare occurrence in the natural history of type 2 diabetes otherwise.^18^ However, DKA was previously identified be uncommon among CAM compared to mucormycosis not associated with COVID-19 (8.6% vs 27%, p<0.001).^1^ Additionally, we found progressively higher risk of mucormycosis with higher blood glucose levels suggesting a dose-response relationship, with those having blood glucose >400 mg/dL, at the highest risk.

Prolonged corticosteroid intake is a risk factor for the development of mucormycosis.^15^ This may be mediated by inhibition of macrophages and neutrophils and tendency to cause hyperglycemia. Steroid-induced hyperglycemia (SIH) primarily raises post-prandial glucose levels and their measurement is imperative for glycemic control. High dose steroid use for short term does not raise HbA1c in those without diabetes or prediabetes, and higher pre-steroid therapy HbA1c predicts development of SIH.^19^ We found that patients in the CAM group had 7.7 [95%CI 2.4–24.7] times higher odds of being administered steroids when compared to the control group. However, if prescribed, the cumulative dose of steroids was similar in both groups. This was probably related to the well-defined regimens for steroid administration in COVID-19. The absolute risk of CAM among patients recovering from COVID-19 is low and should not prevent appropriate prescription of steroids in moderate or severe COVID-19 due to their proven benefit. ^20^ Our findings call for avoiding use of steroids in mild COVID-19 in view of the risk of CAM. Overall, it appears that neutrophil dysfunction (due to COVID-19, hyperglycemia, and steroids) and endothelial dysfunction (due to diabetes and COVID-19) may be the main pathogenetic mediators of CAM.

Although studies on zinc in COVID-19 have shown equivocal results, it is widely used owing to its good safety profile and low cost.^21^ Zinc is essential for normal functioning of immune cells in our body. Further, zinc supplementation has been hypothesized to reduce proinflammatory cytokines, blunt lymphopenia and reduce airway inflammation in COVID-19 patients.^22^ We found that zinc supplementation was associated with significant reduction in the risk of mucormycosis, but a dose-response relationship was not seen. This may be related to the beneficial immuno-modulatory role of zinc, especially in COVID-19 patients who are likely to be deficient.^23^ Prospective clinical trials are needed to confirm these findings.

### Role of COVID-19 disease severity including ferritin

More than one-third of patients in the CAM arm (37.7%) had recovered from moderate to severe COVID-19. This is in contrast to the COSMIC study in which 72% patients recovered from moderate or severe COVID-19 disease.^9^ Our results suggest that severe disease was associated with a lower risk of developing mucormycosis. This may be related the selection bias of recruiting mainly hospitalized patients in both arms, which may have enriched the control arm with patients having severe disease. Secondly, survivorship bias may have played a role since patients of severe COVID-19 may have died prior to development of symptoms of CAM. A prospective study design is required to evaluate the same.

The overall proportion of patients requiring oxygen therapy as well as the duration of oxygen therapy was similar in cases and controls. However, exposure to high flow oxygen devices was lower in the CAM group in line with the proportion of patients with severe COVID-19 illness.

COVID-19 disease severity correlates with raised ferritin levels reflecting the inflammatory state. Ferritin levels were found to be higher in cases of CAM compared to controls. Iron uptake correlates linearly with growth of *Rhizopus* in serum and is essential for its pathogenicity in the presence of siderophores.^24^ Higher ferritin levels have been associated, albeit weakly, with the risk of mucormycosis in hematopoietic allograft recipients.^25^ The pathogenetic role of iron metabolism in mucormycosis merits further evaluation as elevated ferritin does not directly imply increased intracellular iron available to *Mucorales*.

### Practices related to COVID-19 prevention and diagnosis

*Rhizopus*, the dominant species responsible for mucormycosis, is thermotolerant and grows well at or above body temperature and in a moist environment.^26^ This prompted us to evaluate the role of mask use in modulating the risk of CAM. Use of surgical or cloth masks for prolonged periods was found to be associated with an increased risk of CAM in our study. Temperature of the perioral skin rises after use of use of face masks for even one hour.^27^ Cloth masks increase the risk of respiratory tract infections compared to other masks, as well as no mask use.^28^ The reuse of cloth masks, variation in effective cleaning practices, and retention of moisture are proposed as factors for this increase. N95 masks are made of fluid resistant synthetic materials that inhibit the growth of fungal elements.^29^ Organic cloths used commonly in Indian masks, like cotton and silk, do not share similar properties. The organic matter contaminating usual cloth masks may allow implantation of *Mucorales* spores within the nasal cavity. Also, duration of mask use may be a surrogate for increased outdoor exposure and consequently soil and dust that harbor *Mucorales* spores. Our findings may require further evaluation in prospective studies since a large proportion of our controls used N95 masks (42.5% vs 18% in CAM), probably due to the disproportionate number of healthcare workers in that arm (n=40). We also could not obtain reliable information about frequency of discarding or laundering of masks, socio-cultural factors influencing the preferred mask type, or change in mask use practices during recovery from COVID-19 – at a time when CAM is in incubation. Nevertheless, the use of unclean cloth or surgical masks should be avoided.

Physical factors that may increase the risk of mucormycosis were also evaluated. Repeated nasopharyngeal swab testing for COVID-19, if done more than twice, was found to be independently associated with higher risk of CAM. Development of cutaneous mucormycosis at sites of local trauma such as infected intravenous catheters, dressings and needles is well reported.^30^ Repeated nasopharyngeal swab testing may cause microtrauma to nasal and nasopharyngeal mucosa, predisposing to angioinvasion. Similarly, steam inhalation has been hypothesized to cause heat injury to the nasal mucosa, whereas we did not find any association between the risk of CAM and frequent steam use or continued steam use beyond recovery from COVID-19.^31^

Our study is the first large case-control study evaluating the risk factors specific to development of CAM. We hypothesized and investigated several risk factors for CAM unique to the prevention and management of COVID-19, beyond those already established for mucormycosis prior to the COVID-19 pandemic. Information was obtained and corroborated from several sources to mitigate recall bias and missing information. Restricting ourselves to the second wave of COVID-19 between April-May 2021 prevented discrepancies in healthcare access due to the overwhelming rise in cases. The peak incidence of COVID-19 symptoms for both groups occurred on 20^th^ April 2021 (Figure 1), coinciding with the peak number of 28,395 COVID-19 cases reported in a single day in the region of Delhi, India as per crowd-sourced trackers.^32^ However, this study has a few limitations. Firstly, recall bias is inherent to the case-control study design and cannot be completely eliminated. Secondly, difference in the standard of care of COVID-19 treatment practices may exist since CAM cases were treated at other hospitals while controls were mainly treated at our tertiary care center. Thirdly, since it was a retrospective study, the robustness of our findings decreased due to lower sample size available for multivariable modelling in view of missing information particularly for blood glucose, HbA1c and ferritin. Fourth, cases of CAM may occur infrequently beyond 30 days of COVID-19 onset (n=10, 6.6%) and thus controls may remain at-risk of CAM. However, none of the controls have notified the authors of the occurrence of CAM even after study completion.

In conclusion, CAM is strongly associated with diabetes, poor glycemic control, and systemic steroid use. Requirement of oxygen therapy and hospitalization for COVID-19 did not affect the risk of CAM. Novel risk factors identified in our study include prolonged use of cloth and surgical masks vis-a-vis N95 masks, and repeated nasopharyngeal swab testing. These are potentially modifiable and merit further prospective research.

## Data Availability

Data will be available upon request to the corresponding author, along with a proposed research question explaining the request for data.

## Acknowledgements

None

## Funding source

None to declare

## Notes

### Competing Interest Statement

The authors have declared no competing interest.

### Author Declarations

The study was approved by the Institute Ethics Committee of AIIMS, Ansari Nagar, Delhi, India on 28/05/2021. The reference number for the approval is IECPG-353/28.05.21.

## References

1. Patel A, Agarwal R, Rudramurthy SM, Shevkani M, Xess I, Sharma R, et al. Multicenter Epidemiologic Study of Coronavirus Disease–Associated Mucormycosis, India. Emerg Infect Dis. 2021 Sep;27(Early Release).

2. Chakrabarti A, Dhaliwal M. Epidemiology of Mucormycosis in India. Curr Fungal Infect Rep. 2013 Dec 1;7(4):287–92.

3. Hindustan Times. Black fungus: Here is a list of states with highest number of mucormycosis cases [Internet]. Hindustan Times. 2021 [cited 2021 Jun 21]. Available from: https://www.hindustantimes.com/india-news/black-fungus-states-with-highest-number-of-mucormycosis-cases-101621559394002.html

4. Cornely OA, Alastruey-Izquierdo A, Arenz D, Chen SCA, Dannaoui E, Hochhegger B, et al. Global guideline for the diagnosis and management of mucormycosis: an initiative of the European Confederation of Medical Mycology in cooperation with the Mycoses Study Group Education and Research Consortium. Lancet Infect Dis. 2019 Dec;19(12):e405–21.

5. WHO COVID-19 Case definition [Internet]. [cited 2021 Jun 21]. Available from: https://www.who.int/publications-detail-redirect/WHO-2019-nCoV-Surveillance_Case_Definition-2020.2

6. Liu D, Ahmet A, Ward L, Krishnamoorthy P, Mandelcorn ED, Leigh R, et al. A practical guide to the monitoring and management of the complications of systemic corticosteroid therapy. Allergy Asthma Clin Immunol. 2013 Dec;9(1):1–25.

7. Patel A, Kaur H, Xess I, Michael JS, Savio J, Rudramurthy S, et al. A multicentre observational study on the epidemiology, risk factors, management and outcomes of mucormycosis in India. Clin Microbiol Infect. 2020 Jul;26(7):944.e9-944.e15.

8. Chakrabarti A, Singh R. Mucormycosis in India: unique features. Mycoses. 2014 Dec;57:85–90.

9. Sen M, Honavar SG, Bansal R, Sengupta S, Rao R, Kim U, et al. Epidemiology, clinical profile, management, and outcome of COVID-19-associated rhino-orbital-cerebral mucormycosis in 2826 patients in India – Collaborative OPAI-IJO Study on Mucormycosis in COVID-19 (COSMIC), Report 1. Indian J Ophthalmol. 2021 Jul;69(7):1670–92.

10. Ghuman H, Voelz K. Innate and Adaptive Immunity to Mucorales. J Fungi Basel Switz. 2017 Sep 5;3(3):E48.

11. Reusch N, De Domenico E, Bonaguro L, Schulte-Schrepping J, Baßler K, Schultze JL, et al. Neutrophils in COVID-19. Front Immunol. 2021 Mar 25;12:652470.

12. Lucas C, Wong P, Klein J, Castro TBR, Silva J, Sundaram M, et al. Longitudinal analyses reveal immunological misfiring in severe COVID-19. Nature. 2020 Aug;584(7821):463–9.

13. Jin Y, Ji W, Yang H, Chen S, Zhang W, Duan G. Endothelial activation and dysfunction in COVID-19: from basic mechanisms to potential therapeutic approaches. Signal Transduct Target Ther. 2020 Dec 24;5(1):293.

14. Afiat B, Nofri R, Adi IT, Rovina R. Type 2 Diabetes and its Impact on the Immune System. Curr Diabetes Rev. 2020 May 31;16(5):442–9.

15. Ibrahim AS, Spellberg B, Walsh TJ, Kontoyiannis DP. Pathogenesis of Mucormycosis. Clin Infect Dis. 2012 Feb 1;54(suppl_1):S16–22.

16. Binder U, Maurer E, Lass-Flörl C. Mucormycosis--from the pathogens to the disease. Clin Microbiol Infect Off Publ Eur Soc Clin Microbiol Infect Dis. 2014 Jun;20 Suppl 6:60–6.

17. Unsworth R, Wallace S, Oliver NS, Yeung S, Kshirsagar A, Naidu H, et al. New-Onset Type 1 Diabetes in Children During COVID-19: Multicenter Regional Findings in the U.K. Diabetes Care. 2020 Nov;43(11):e170–1.

18. Dhatariya KK, Glaser NS, Codner E, Umpierrez GE. Diabetic ketoacidosis. Nat Rev Dis Primer. 2020 May 14;6(1):40.

19. Vidler J, Rogers C, Yallop D, Devereux S, Wellving E, Stewart O, et al. Outpatient management of steroid-induced hyperglycaemia and steroid-induced diabetes in people with lymphoproliferative disorders treated with intermittent high dose steroids. J Clin Transl Endocrinol. 2017 Sep;9:18–20.

20. The RECOVERY Collaborative Group. Dexamethasone in Hospitalized Patients with Covid-19. N Engl J Med. 2021 Feb 25;384(8):693–704.

21. Thomas S, Patel D, Bittel B, Wolski K, Wang Q, Kumar A, et al. Effect of High-Dose Zinc and Ascorbic Acid Supplementation vs Usual Care on Symptom Length and Reduction Among Ambulatory Patients With SARS-CoV-2 Infection: The COVID A to Z Randomized Clinical Trial. JAMA Netw Open. 2021 Feb 12;4(2):e210369.

22. Corrao S, Mallaci Bocchio R, Lo Monaco M, Natoli G, Cavezzi A, Troiani E, et al. Does Evidence Exist to Blunt Inflammatory Response by Nutraceutical Supplementation during COVID-19 Pandemic? An Overview of Systematic Reviews of Vitamin D, Vitamin C, Melatonin, and Zinc. Nutrients. 2021 Apr 12;13(4):1261.

23. Jayawardena R, Sooriyaarachchi P, Chourdakis M, Jeewandara C, Ranasinghe P. Enhancing immunity in viral infections, with special emphasis on COVID-19: A review. Diabetes Metab Syndr Clin Res Rev. 2020 Jul;14(4):367–82.

24. Boelaert JR, de Locht M, Van Cutsem J, Kerrels V, Cantinieaux B, Verdonck A, et al. Mucormycosis during deferoxamine therapy is a siderophore-mediated infection. In vitro and in vivo animal studies. J Clin Invest. 1993 May;91(5):1979–86.

25. Dadwal SS, Tegtmeier B, Liu X, Frankel P, Ito J, Forman SJ, et al. Impact of pretransplant serum ferritin level on risk of invasive mold infection after allogeneic hematopoietic stem cell transplantation. Eur J Haematol. 2015 Mar;94(3):235–42.

26. Ribes JA, Vanover-Sams CL, Baker DJ. Zygomycetes in human disease. Clin Microbiol Rev. 2000 Apr;13(2):236–301.

27. Scarano A, Inchingolo F, Lorusso F. Facial Skin Temperature and Discomfort When Wearing Protective Face Masks: Thermal Infrared Imaging Evaluation and Hands Moving the Mask. Int J Environ Res Public Health. 2020 Jun 27;17(13):E4624.

28. MacIntyre CR, Seale H, Dung TC, Hien NT, Nga PT, Chughtai AA, et al. A cluster randomised trial of cloth masks compared with medical masks in healthcare workers. BMJ Open. 2015 Apr 1;5(4):e006577.

29. Bhattacharjee S, Bahl P, Chughtai AA, MacIntyre CR. Last-resort strategies during mask shortages: optimal design features of cloth masks and decontamination of disposable masks during the COVID-19 pandemic. BMJ Open Respir Res. 2020 Sep;7(1):e000698.

30. Jeong W, Keighley C, Wolfe R, Lee WL, Slavin MA, Kong DCM, et al. The epidemiology and clinical manifestations of mucormycosis: a systematic review and meta-analysis of case reports. Clin Microbiol Infect. 2019 Jan;25(1):26–34.

31. Banerjee M, Pal R, Bhadada SK. Intercepting the deadly trinity of mucormycosis, diabetes and COVID-19 in India. Postgrad Med J [Internet]. 2021 Jun 7 [cited 2021 Jul 23]; Available from: https://pmj.bmj.com/content/early/2021/06/08/postgradmedj-2021-140537

32. Covid19india.org. Coronavirus in India [Internet]. [cited 2021 Jun 21]. Available from: https://www.covid19india.org

